# Multispectral Fundus Photography of Choroidal Nevi with Trans-Palpebral Illumination

**DOI:** 10.1101/2024.01.12.24301119

**Authors:** Mojtaba Rahimi, Alfa Rossi, Taeyoon Son, Albert K. Dadzie, Behrouz Ebrahimi, Mansour Abtahi, Michael J. Heiferman, Xincheng Yao

**Affiliations:** Department of Biomedical Engineering, University of Illinois Chicago, Chicago, IL 60607, USA; Department of Ophthalmology and Visual Sciences, University of Illinois Chicago, Chicago, IL, 60612, USA

## Abstract

**Purpose:** To investigate the spectral characteristics of choroidal nevi and assess the feasibility of quantifying the basal diameter of choroidal nevi using multispectral fundus images captured with trans-palpebral illumination.

**Methods:** The study employed a widefield fundus camera with multispectral (625 nm, 780 nm, 850 nm, and 970 nm) trans-palpebral illumination. Geometric features of choroidal nevi, including border clarity, overlying drusen, and lesion basal diameter, were characterized. Clinical imagers, including scanning laser ophthalmoscopy (SLO), autofluorescence (AF), and optical coherence tomography (OCT), were utilized for comparative assessment.

**Results:** Fundus images captured with trans-palpebral illumination depicted nevi as dark regions with high contrast against the background. Near-infrared (NIR) fundus images provided enhanced visibility of lesion borders compared to visible light fundus images and SLO images. Lesion-background contrast measurements revealed 635 nm SLO at 11% and 625 nm fundus at 42%. Significantly enhanced contrasts were observed in NIR fundus images at 780 nm (73%), 850 nm (63%), and 970 nm (67%). For quantifying the basal diameter of nevi, NIR fundus images at 780 nm and 850 nm yielded a deviation of less than 10% when compared to OCT B-scan measurements.

**Conclusion:** NIR fundus photography with trans-palpebral illumination enhances nevi visibility and boundary definition compared to SLO. Agreement in basal diameter measurements with OCT validates the accuracy and reliability of this method for choroidal nevi assessment.

**Translational Relevance:** Multispectral fundus imaging with trans-palpebral illumination improves choroidal nevi visibility, accurately measures basal diameter, promising to enhance clinical practices in screening, diagnosis, and monitoring of choroidal nevi.

## Introduction

Choroidal nevus is the most frequently occurring primary intraocular tumor^1^. This condition poses two associated risks: firstly, a benign nevus can transform into malignant melanoma, with risks of melanoma-related metastasis and mortality; and secondly, nevi may lead to vision impairment from subretinal fluid or secondary choroidal neovascularization^2, 3^. Early detection and careful progression monitoring of choroidal nevi are crucial as they are early-stage precursors to choroidal melanomas^4^. Shields et al.^5^ have proposed several risk factors associated with the transformation of choroidal nevus into melanoma. These factors include lesion thickness, the presence of subretinal fluid, visual symptoms, the presence of orange pigment, and ultrasound acoustic hollowness. Clinicians rely on various imaging modalities, including color fundus photography (CFP)^6^, scanning laser ophthalmoscopy (SLO)^7^, fundus autofluorescence (AF)^8^, optical coherence tomography (OCT)^9, 10^, and ultrasonography^11^, to assess these risk factors. Each of these imaging modalities has its own strengths and limitations. Consequently, multimodal imaging is often employed for the assessment of choroidal nevi^5^. CFP and SLO provide two-dimensional lateral profiles of the lesion. In contrast, OCT and ultrasound images reveal depth information of nevi. Additionally, AF image can reveal the accumulation of lipofuscin, seen clinically as overlying orange pigment, suggestive of tumor activity. Ultrasonography is currently the gold standard for the measurement of intraocular tumors and provides an accurate measurement of tumor thickness using A-scan. However, it is acknowledged that B-scan ultrasonography measurements of lateral diameter are less precise for choroidal tumors, especially for choroidal nevi that appear flat due to their thinness^7^. Nevertheless, small choroidal tumors that might escape detection via ultrasonography can be objectively measured using OCT which provides high-resolution cross sectional imaging of choroidal lesions^10, 12^. However, OCT is still relatively expensive, requires pharmacologic dilation, and can be inaccessible for patients in rural and underserved areas where advanced imaging equipment and skilled operators are limited.

Conventional CFP is employed for imaging choroidal lesions due to its availability and cost-efficiency. However, it presents two significant limitations for the imaging and assessment of choroidal nevi: a limited field of view (FOV) and low choroidal lesion contrast. FOV of traditional fundus cameras typically ranges from 30° to 45° visual angle (45° to 68° eye angle) as it requires careful allocation of the available pupil for both illuminating and imaging pathes^13, 14^. Consequently, it becomes challenging to detect choroidal nevi, especially those located in the periphery of the fundus. Ultra-widefield SLO imaging systems (e.g., Optos), offering a 134° visual angle (200° eye angle) FOV^15^, serve as an alternative means to image choroidal nevi and measure tumor basal diameter^7^. However, SLO imagers incorporate several laser sources and scanning systems that increase the cost and complexity of the system.

Trans-palpebral illumination has been investigated as an alternative illumination method in fundus photography, enabling widefield fundus imaging without the need for pupil dilation^16–18^. Son et al^17^. achieved a 93° visual angle (140° eye angle) FOV in a single snapshot fundus image utilizing trans-palpebral illumination. The incorporation of narrow-band light emitting diodes (LEDs) into this method has demonstrated the capability to enable multispectral imaging of the retina and choroid^17^. In contrast to traditional fundus photography which relies on white light illumination to capture a color fundus image by combining reflection information from several retinal layers^19, 20^, multispectral fundus photography selectively image different layers of retina and choroid^21^. Generally, longer wavelengths of light allow deeper penetration through the retina into the choroid (Figure 1 (B)). Therefore, short visible light predominantly uncovers superficial retinal structure, while long visible wavelength and near-infrared (NIR) light images are attributed to both the retinal and choroidal layers^22^. Choroidal nevi originate within the choroid beneath the retina and can impact the overlying retinal pigment epithelium (RPE), resulting in features like atrophy, drusen, and lipofuscin accumulation^23^. As a result, multispectral fundus photography using long visible and NIR wavelengths promises a practical solution for selectively capturing these choroidal lesions. However, most multispectral fundus devices are essentially conventional fundus cameras that incorporate multiple light sources or tunable filters into their trans-pupillary illumination^24–26^. Consequently, they tend to have bulky dimensions, and spatial scanning is required due to the limited FOV.

**Figure 1.**
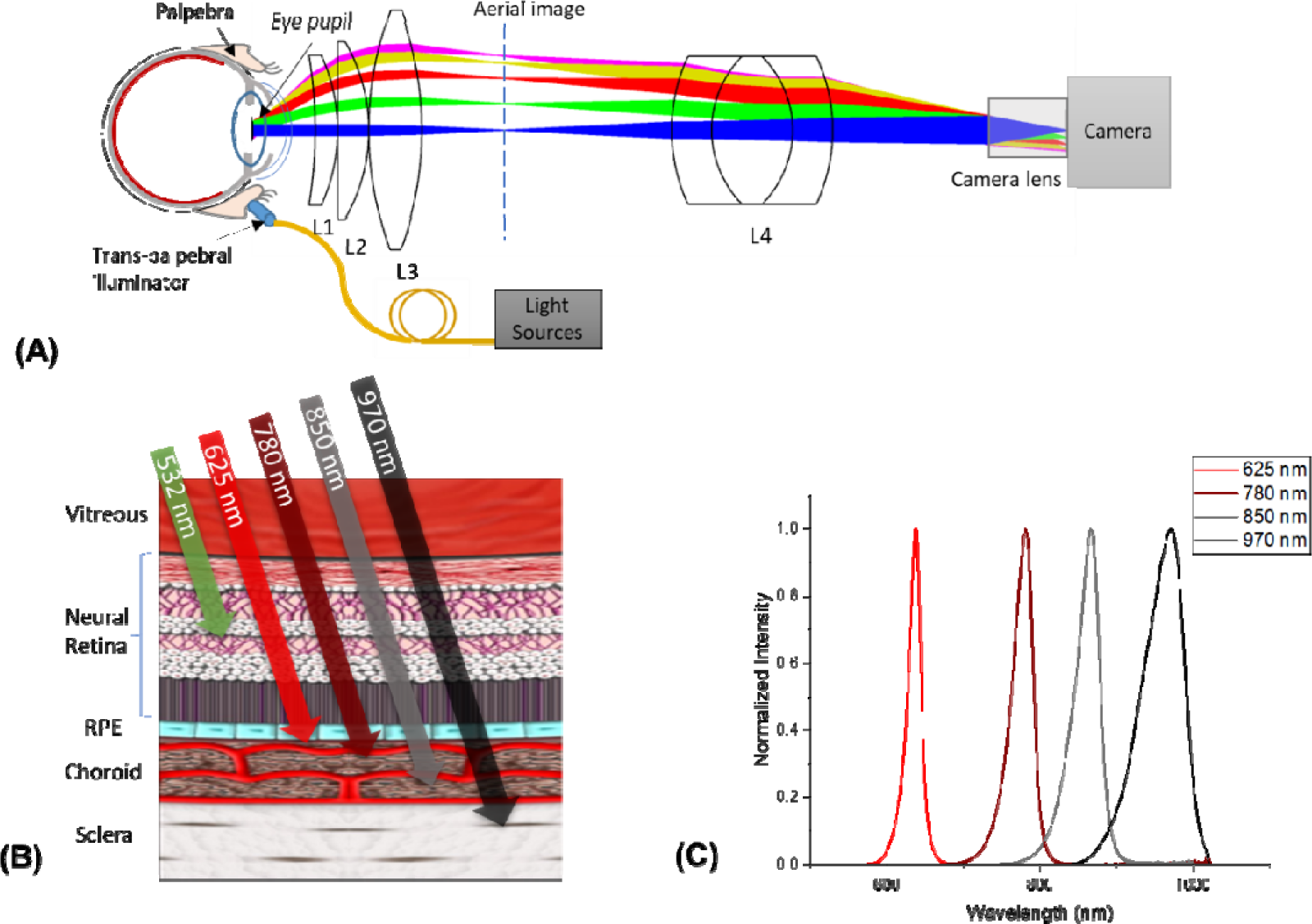
A) Optical layout of the system with trans-palpebral illumination, B) Light penetration in the retina and choroid (modified from 35), C) visible and NIR LED spectra.

Previous studies by Muftuoglu et al.^6^ and Venkatesh, et al^27^. have compared the effectiveness of multispectral SLO imaging to CFP in identifying choroidal lesions. They reported that multispectral SLO provides similar information with CFP for the features of choroidal lesions. Moreover, Azzolini et al.^28^ demonstrated that the retro-mode and dark-field mode of SLO can enhance the contrast of choroidal nevi and delineate lesion margins in NIR wavelength. However, this enhancement necessitates specific adjustments to the confocal laser scanning system. On the other hand, trans-palpebral illumination naturally fulfills the requirement for a dark-field imaging condition, enhancing the contrast of choroidal lesions. In this oblique illumination method, light is delivered from a region outside of the pupil, typically with a ∼4 mm distance from the limbus. This contrast enhancement is valuable for detecting and monitoring the basal diameter of choroidal nevi. Furthermore, the cost-effectiveness of multispectral fundus photography with trans-palpebral illumination, along with the absence of the need for pupillary dilation, indicates its potential as an optimal choice for screening the general population for choroidal nevi. Choroidal nevi affect approximately 5% of the population and are recommended to be monitored at least once per year, given that roughly one-third of uveal melanomas are asymptomatic^3, 29^. Therefore, this method can empower referring healthcare professionals, such as primary care physicians and optometrists, to screen and triage patients more effectively based on geometric features of the lesion. This study aims to investigate the effectiveness of multispectral fundus photography with trans-palpebral illumination as an affordable approach for high contrast imaging and quantitative assessment of choroidal nevi.

## Methods

### Experimental setup

Figure 1 (A) illustrates the optical configuration of the widefield fundus camera system, including trans-palpebral illumination for multispectral fundus images. The first lens group (L1, L2, and L3) consists of a meniscus lens (L1: LE1234-A, Thorlabs Inc., Newton, NJ), a plano-convex lens (L2: 67–152, Edmund Optics Inc., Barrington, NJ), and a double-convex lens (L3: 63–688, Edmund Optics Inc., Barrington, NJ, USA). This lens assembly collects light from the pupil and generates an aerial image in front of the relay optics. Subsequently, a triplet achromatic lens (L4: 67–422, Edmund Optics Inc., Barrington, NJ, USA) and the camera lens (33–303, Edmund Optics Inc., Barrington, NJ, USA) are precisely positioned to relay this aerial image onto the camera sensor. The removal of the infrared filter from the color CMOS camera (DFK 37BUX250, The Imaging Source Europe GmbH, Bremen, Germany) enables the detection of NIR light. By delivering light through the eyelid, the trans-palpebral illuminator provides a convenient means of illuminating the interior of the eye. It comprises 1-to-4 fan-out fiber bundles (BF46LS01, Thorlabs Inc., Newton, NJ). Each individual optical fiber within these bundles possesses a diameter of 600 µm and a numerical aperture of 0.39. These optical fibers are connected to narrow-band LED light sources with center wavelengths of 625 nm (M625L4, Thorlabs Inc., Newton, NJ), 780 nm (M780L3, Thorlabs Inc., Newton, NJ), 850 nm (M850LP1, Thorlabs Inc., Newton, NJ), and 970 nm (M970L4, Thorlabs Inc., Newton, NJ). The optical spectrum of the light sources is shown in Figure 1 (C). The specifics of the optical system design have been elaborated upon in previous publication^17^.

### Human subjects and imaging procedures

This study received approval from the Institutional Review Board at the University of Illinois Chicago and adhered to the ethical principles outlined in the Declaration of Helsinki. It included eight eyes from eight patients previously diagnosed with a choroidal nevus. Informed consent was obtained from each subject prior to imaging. All participants underwent comprehensive clinical multimodal imaging procedures, which included ultra-widefield SLO, fundus AF, NIR reflectance, and spectral domain OCT enabling a comparative analysis. OCT and NIR reflectance were performed using Spectralis (Heidelberg Engineering GmbH; Heidelberg, Germany). SLO and AF images were captured using Optos California (Optos; Marlborough, MA, USA). Furthermore, multispectral fundus images were obtained using the fundus camera equipped with trans-palpebral illumination. For image stability, participants were positioned with their foreheads and chins resting on supports. The trans-palpebral illuminator was gently positioned on the eyelid and individually adjusted based on the live preview image. A custom LabVIEW interface was used to enable real-time monitoring, facilitating the alignment of the fundus camera and trans-palpebral illuminator, as well as the sequential capture of fundus images. To aid in maintaining fixation during the imaging procedure, a low-intensity LED served as a visual target.

### Light safety

The ISO standard for “Ophthalmic Instrument-Fundus camera”^30^ is employed to evaluate ocular light safety for multispectral fundus imaging with trans-palpebral illumination, considering both photochemical and thermal hazards. The LED powers at the output of the fiber were adjusted to 15 mW, 10 mW, 7 mW, and 5 mW for 625 nm, 780 nm, 850 nm, and 970 nm illuminations, respectively. According to the ISO standard, to prevent thermal hazards, the highest allowable weighted power intensity on the retina is 700 mW/cm^2^. The methodology for calculating the illuminated retinal area during trans-palpebral illumination was explained in a previous publication^31^. Following the assessment of thermal hazards, the calculated equivalent power levels for the light sources at 625 nm, 780 nm, 850 nm, and 970 nm were determined to be 13.1 mW/cm^2^, 15.9 mW/cm^2^, 14.4 mW/cm^2^, and 7.9 mW/cm^2^, respectively. The calculated weighted power intensity for all of these wavelengths was found to be significantly lower than the established allowable limit, indicating that there were no thermal hazard concerns associated with any of the considered wavelengths. The photochemical hazard is negligible for NIR wavelengths. The weighted irradiance for the visible light was computed utilizing the photochemical hazard weighing function specified in the ISO standard. For 625 nm illumination, the aphakic photochemical hazard weighing function is determined to be 0.001. Consequently, the calculated maximum permissible exposure time exceeds 24 hours. A comprehensive description of the ocular safety calculations can be found in a prior publication^32^.

## Results

### Lateral Characterization

Figure 2 represents the results of imaging a participant diagnosed with a choroidal nevus. In Figure 2 (A1) and (A2), the SLO image and an enlarged view of the nevus region are presented, respectively. Figure 2 (B1) to (B4) shows the green and red channels of the SLO image, the AF image, and the NIR reflectance images. Representative multispectral fundus images with 625 nm, 780 nm, 850 nm, and 970 nm trans-palpebral illumination are shown in Figure 2 (C1) to Figure 2 (C4), respectively. For direct comparison, the green and red channels of the SLO image and AF image have been cropped to correspond to the same region depicted in the fundus images. The green channel of SLO (Figure 2 (B1)), generated through a 532 nm laser, provides insight into the superficial structures of the retina to the RPE layer. In contrast, the red channel (Figure 2 (B2)), produced with a 635 nm laser, predominantly accentuates the deeper retinal structures, encompassing the RPE through the choroid. The AF image (Figure 2 (B3)) reveals the minimal fluorescent properties at the RPE layer. An 815 nm laser is used to capture the NIR reflectance image (Figure 2 (B4)). The fundus image captured using visible light (Figure 2 (C1)) revels retinal veins and some choroidal structure due to the partial penetration of red light through the RPE layer. When employing 780 nm illumination (Figure 2 (C2)), an improved depiction of the choroidal vasculature is achieved compared to red illumination. This aligns with the concept of reduced melanin absorption in the RPE, consequently allowing enhanced NIR light penetration to reveal the choroidal vasculature. By extending the illumination wavelength to 970 nm (Figure 2 (C4)), predominantly choroidal veins became discernible in the fundus image.

**Figure 2.**
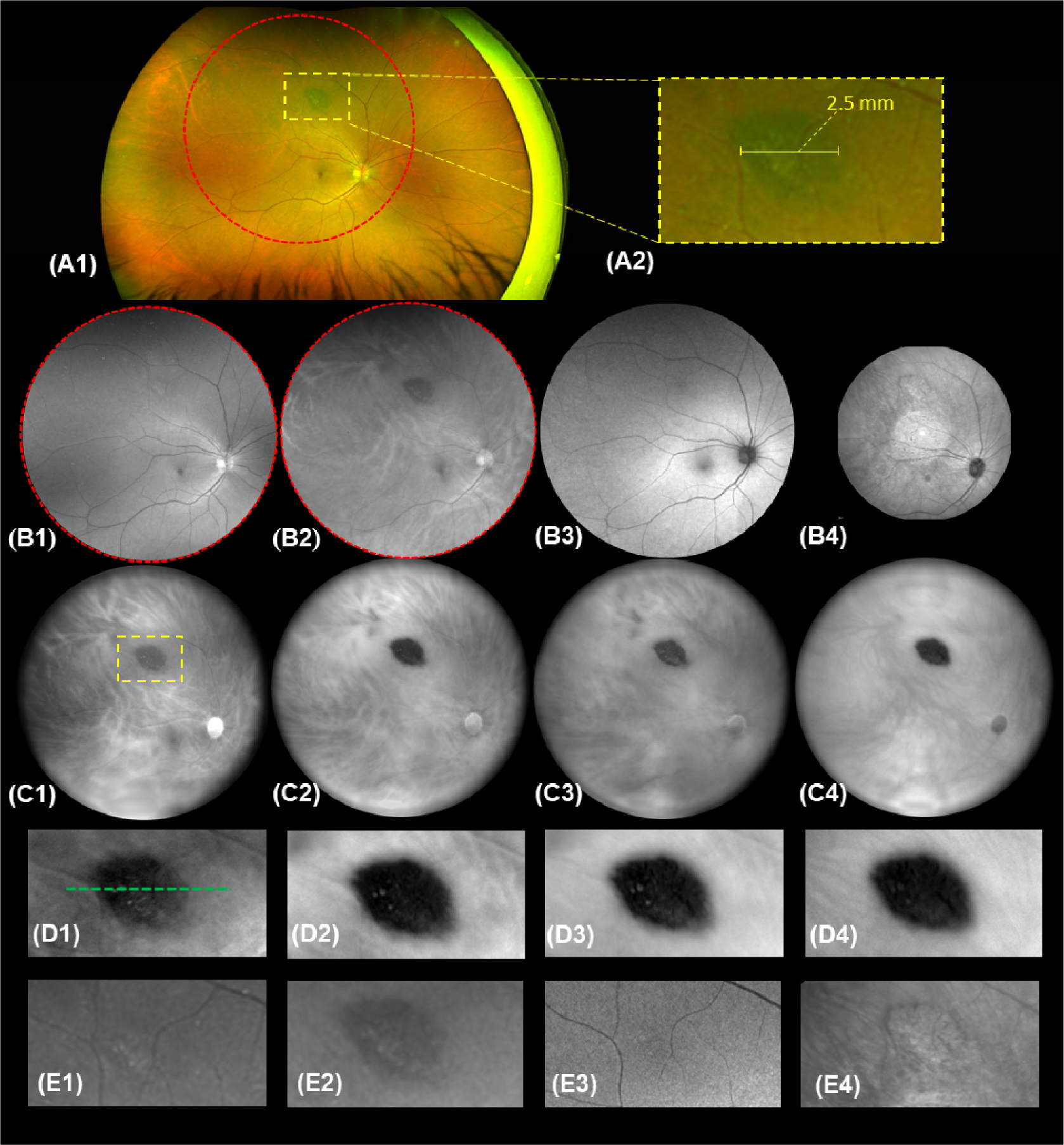
A) Representative images of a choroidal nevus, A1) SLO image, A2) enlarged nevus region, B1) Cropped green channel of SLO corresponding to red-dashed circle in the SLO image, B2) red channel of SLO, B3) AF image, B4) NIR reflectance image, C) Multispectral fundus images captured using C1) 625 nm, C2) 780 nm, C3) 850 nm, and C4) 970 nm trans-palpebral illumination, D) Enlarged nevus region in the fundus images captured using D1) 625 nm, D2) 780 nm, D3) 850 nm, and D4) 970 nm trans-palpebral illumination, enlarged nevus region in E1) green channel and E2) red channel of SLO, E3) AF, and E4) NIR reflectance images.

To facilitate a more straightforward comparison of nevi using various imaging modalities, Figure 2 (D) presents the enlarged nevus region within the multispectral fundus images captured under trans-palpebral illumination. Additionally, Figure 2 (E) displays the enlarged nevus region within the green and red channels of the SLO (Figure 2 (E1 and E2)), AF (Figure 2 (E3)), and NIR (Figure 2 (E4)) images. The nevus is observed as a distinct dark region in both the red (Figure 2 (D1)) and NIR (Figure 2 (D2-D4)) fundus images, significantly more visible in comparison to the SLO image (Figure 2 (E2)) and the AF image (Figure 2 (E3)). Additionally, the NIR fundus images exhibited an improved capacity to illustrate clear boundaries of the nevus, as well as to highlight information about overlying drusen. Analysis of the green and red channels of the SLO image indicates that the green channel (Figure 2 (E1)) does not reveal the nevus, while the nevus appears as a hypo-reflective region in the red channel (Figure 2 (E2)), though its boundaries remain unclear. Furthermore, the nevus does not exhibit significant autofluorescence features in the AF image ((Figure 2 (E3))) due to absence of overlying orange pigment. However, the overlying drusen can be seen faintly in the AF image. In the NIR reflectance image (Figure 2 (E4)), the nevus is characterized by an iso-reflective region with overlying drusen. The diameter of the nevus is measured utilizing the measurement tool in the Optos-Advance software (Figure 2 (A2)). The diameter measurements in the SLO image rely on the color contrast of the lesion within the image, a method described by Pe’er et al^7^.

To evaluate the measurement of choroidal nevus basal diameter in fundus images, the intensity line profiles derived from fundus images captured under visible and NIR illumination are analyzed. The intensity line profile of the lesion, indicated by a dashed line in Figure 2 (D1), is presented in Figure 3 (A). The full width at half maximum (FWHM) of the normalized intensity profile serves as the reference for measurement of nevus basal diameter in fundus images. Furthermore, Figure 3 (B) displays the intensity profile of the same region in the green and red channels of SLO, AF, and NIR reflectance images, showing the presence of overlying drusen in all intensity profiles. Analyzing the intensity profile of the nevus in the green and red channels of SLO reveals that the contrast of the lesion compared to the surrounding region originates from the red channel. This contrast is somewhat less pronounced than the contrast observed in the fundus images. Lesion-background mean contrast was considered to quantify the visibility of the nevus in the images. The straightforward definition of Lesion-background contrast (C) is as follows:^33^

**Figure 3.**
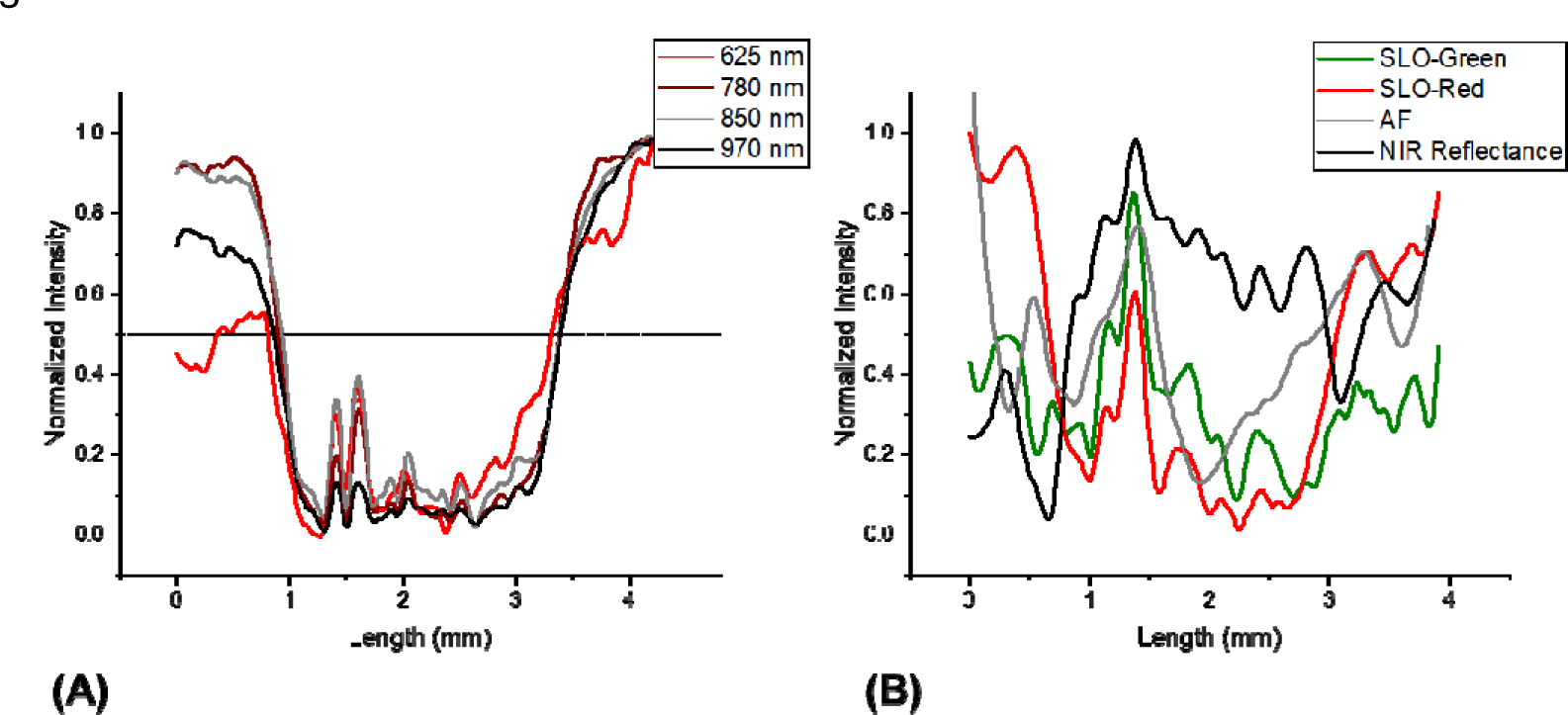
A) The intensity profile of the nevus in the fundus images along the green dashed line in Figure 2 (D1), B) the intensity profile of the nevus in the green and red channels of SLO, AF, and NIR reflectance images.

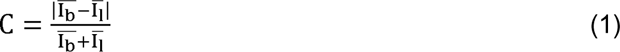

where I^-^ and I^-^ are the mean intensity of the lesion and its background, respectively. The red channel of the SLO image and the 625 nm fundus image exhibited lesion-background contrasts of 11% and 42%, respectively. However, fundus images with NIR illumination at 780 nm, 850 nm, and 970 nm showed higher lesion-background contrasts, measuring 73%, 63%, and 67%, respectively. The results of nevus basal diameter measurement and quantification of lesion-background contrasts for the other seven subjects are summarized in Table 1. The consistency of the results across all subjects supports the validity of the study.

**Table 1.** Characteristics of participant subjects and comparative analysis: Nevus basal diameter measurement in OCT B-Scan vs. widefield fundus images with NIR illumination, and nevus contrast comparison between multispectral fundus images and the SLO image.

| Subject | Eye | Sex | Nevus diameter ($\mu\text{m}$) | | | | Nevus Contrast (%) | | | | |
| --- | --- | --- | --- | --- | --- | --- | --- | --- | --- | --- | --- |
|  |  |  | OCT scan | Fundus 780 nm | Fundus 850 nm | Fundus 970 nm | SLO-(red) | Fundus 625 nm | Fundus 780 nm | Fundus 850 nm | Fundus 970 nm |
| 1 | OD | F | 2570 | 2475 | 2495 | 2534 | 10.88 | 42.34 | 73.33 | 63.05 | 67.48 |
| 2 | OD | M | 2386 | 2427 | 2430 | 2642 | 8.79 | 22.69 | 66.26 | 65.37 | 60.25 |
| 3 | OD | M | 2691 | 2574 | 2663 | 2722 | 5.59 | - | 59.10 | 50.35 | 54.18 |
| 4 | OS | M | 1847 | 1950 | 1881 | 1861 | 8.57 | 56.31 | 74.81 | 74.24 | 59.21 |
| 5 | OD | F | 1607 | 1591 | 1560 | 1549 | 17.43 | 60.62 | 70.10 | 59.89 | 55.14 |
| 6 | OS | F | 1994 | 1893 | 1799 | 1560 | 22.59 | 42.68 | 56.29 | 53.75 | 36.75 |
| 7 | OD | M | 3054 | 2826 | 2774 | 2722 | 16.26 | 37.04 | 73.40 | 74.42 | 60.83 |
| 8 | OD | F | 1329 | 1256 | 1202 | 1116 | 10.92 | 52.92 | 73.93 | 70.21 | 63.51 |

### Axial Characterization

To characterize the axial property of the choroidal nevus, the OCT B-scan of the nevus region is analyzed. Figure 4 (B) presents the OCT B-scan images of the nevus which exhibits hyperreflectivity along the anterior surface of the lesion, accompanied by dark posterior shadowing. The discernible pattern observed in the choroidal layers of the OCT B-scan greatly aids in measuring the basal diameter of the nevus. To assess the accuracy of quantifying the basal diameter of the nevus in fundus images, the nevus basal diameter was measured using the measurement tool in the Heidelberg Spectralis system within the OCT B-scan (Figure 4 (B)). The dark posterior shadowing observed in the nevus was utilized to define boundaries for diameter measurements in the OCT B-scan, as previously described by Jonna et al^9^. In Table 1, the results of basal diameter measurements in fundus images are compared with those obtained in the OCT B-scan, used as a reference. The results show a deviation of less than 10% in the NIR fundus images at 780 nm and 850 nm compared with measurements obtained from the OCT B-scan, verifying the feasibility and accuracy of quantifying choroidal nevus basal diameter in fundus images. The slight variations in values observed for different wavelengths can be attributed to differences in the nevus shape at various depths and focal planes corresponding to these wavelengths. By increasing the wavelength to 970 nm, the heightened transparency results in reduced contrast and decreased accuracy in measuring lesion diameter. For the third subject listed in Table 1, the 625 nm fundus image exhibited reflection artifacts in the lesion region, rendering the measurements inaccurate. Consequently, this data was excluded from the results.

**Figure 4.**
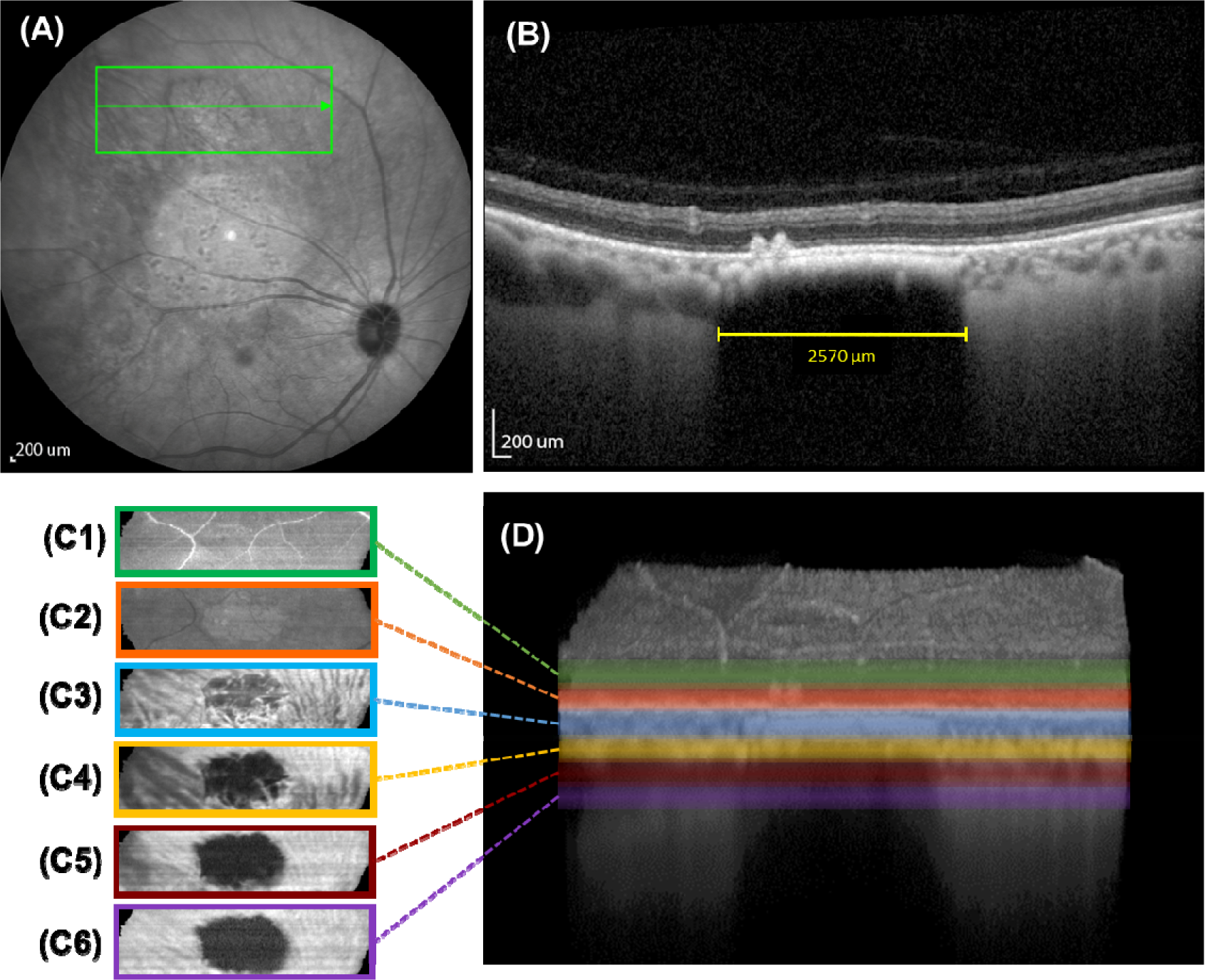
Analysis of OCT scan of the nevus region A) The B-scan line is shown in the NIR reflectance image, B) corresponding OCT B-Scan, C) the mean intensity en face projection maps of OCT B-scans corresponding to highlighted layers in (D), D) OCT image volume of the nevus region after registration and flattening.

In contrast to the cross-sectional B-scan image, the en face OCT image offers a view that resembles traditional 2D imaging. To compare the appearance of the nevus in fundus and OCT en face images, averaged en face OCT images were generated after flattening and registering the OCT B-scans of the nevus region (Figure 4 (C)). In OCT en face images that average retinal layers above the RPE layer (Figure 4 (C1)), the retinal vasculature becomes visible, similar to the green channel of the SLO image (Figure 2 (E1)). In the en face image comprising the RPE layer (Figure 4 (C2)) presence of overlying drusen and RPE alterations can be identified. Additionally, Figure 4 (C3) and Figure 4 (C4) depict the choriocapillaris and medium choroidal vessels of Sattler’s layer, respectively, which exhibit a similar structure to the fundus images captured using 625 nm and 780 nm trans-palpebral illumination (as seen in Figure 2 (D1 and D2)). In Figure 4 (C5), large choroidal vessels in Haller’s layer are visible, and Figure 4 (C6) showcases signals originating from the deep choroid and sclera. These findings correlate with the fundus images captured at longer wavelengths of 850 nm and 970 nm (as shown in Figure 2 (D3 and D4)). The nevus appears as a dark region in OCT en face images from the choroidal layers, resembling the appearance in the fundus image with NIR trans-palpebral illumination. However, the fundus images display overlying drusen, attributable to the fact that longer wavelengths, penetrating deeper layers, still carry information from the layers above.

## Discussion

The spectral characteristics of choroidal nevi and the feasibility of quantifying the basal diameter of these nevi using multispectral fundus images captured with trans-palpebral illumination was investigated. Fundus images captured with the NIR wavelength range revealed choroidal nevi as distinct dark regions with significantly enhanced contrast against the surrounding fundus tissue. The clearer delineation of the nevus boundaries allows clinicians to detect and precisely assess the location and extent of the lesion. Validation of the feasibility of quantifying choroidal nevus diameter in fundus images is achieved by measuring nevus basal diameter in NIR images and comparing it with reference measurements obtained from OCT B-scans.

Previous studies have highlighted the effectiveness of multispectral fundus imaging in selectively capturing detailed images of the retina and choroid^34^. It is widely recognized that choroidal structures can be effectively visualized through the utilization of NIR illumination. According to the study conducted by Burgos-Fernández et al.^26^, choroidal nevi show low reflectance and good contrast at NIR wavelengths in multispectral fundus images. However, their fundus camera utilized trans-pupillary illumination, which restricted the system’s FOV to 30° visual angle (45° eye angle). In addition, Muftuoglu et al.^6^ have compared the multispectral SLO system with CFP. Their multispectral SLO system provided pseudo-color image using three different wavelengths including blue (486 nm), green (518 nm) and NIR (815 nm). Their findings indicated that the multispectral SLO system could offer similar insights to CFP regarding the characteristics of choroidal lesions, including border delineation and the detection of overlying drusen. However, the NIR channel of the multispectral SLO system underestimated the extent of choroidal lesions by approximately 33%. This discrepancy in delineating lesion borders when compared to CFP could be attributed to the confocal imaging system of SLO, which possesses a limited depth of field, in contrast to CFP’s ability to combine reflection data from various depths within the fundus. Furthermore, Azzolini et al.^28^ have shown that utilizing annular and deviated apertures in the confocal plane of SLO provides dark-filed and retro-mode imaging condition in SLO system, which enhanced the contrast of choroidal nevi in NIR SLO image.

In contrast to the SLO system that relies on multiple lasers and mechanical scanning mechanisms, the presented multispectral fundus camera employs LEDs which not only significantly reduce costs but also results in a portable device. These advantages are particularly beneficial for facilitating the emerging field of telemedicine, especially in rural or underserved areas. The fundus camera with trans-palpebral illumination can capture a snapshot with a FOV up to 93° visual angle (140° eye angle) without the need for pupil dilation. By utilizing a fixation target, this FOV can be further expanded to 134° visual angle (200° eye angle)^17^. Furthermore, trans-palpebral illumination benefits from a dark-field imaging condition. This condition is achieved by completely separating the illumination and imaging paths, preventing any specular reflectance light from reaching the camera. The contrast of choroidal nevi within fundus images is primarily a result of heightened light absorption by the melanin and melanocytes present in the nevi^34^. Melanin absorption decreases with increasing wavelength. Consequently, NIR wavelengths can penetrate deeply into the normal choroid and are backscattered from the sclera. The absence of specular reflection from the surface of the nevus in the dark-field imaging condition, along with the backscattered light from the normal choroid tissue, leads to high-contrast in the NIR fundus images with trans-palpebral illumination.

The practical application of trans-palpebral illumination for visible light fundus imaging poses challenges due to the efficiency of light transmission through the eyelid and sclera. It has been demonstrated that this efficiency depends on several factors, including the wavelength of illumination, the location of illuminator, and the complexion of the subject^17^. However, the efficiency of NIR wavelengths in trans-palpebral illumination is significantly higher, by multiple orders of magnitude, compared to visible wavelengths^17^. Consequently, there are no limitations regarding the efficiency of NIR wavelengths for imaging choroidal nevi. Moreover, cutaneous pigmentation effect can be easily compensated for by adjusting the power of the light source, without concerns about light safety. Additionally, an analysis of the spatial dependence of spectral efficiency has shown that the location of illumination significantly affects the efficiency of visible light, while NIR light demonstrates no significant location dependency^31^. These findings along with the affordability and the absence of the need for pupil dilation strongly support the notion that fundus imaging with NIR trans-palpebral illumination offers a straightforward and practical approach for detection and screening of choroidal nevi in the general population. This method has the potential to enable healthcare professionals to assess choroidal nevi more effectively and to expedite referrals to ocular oncologists, ensuring that patients receive timely and appropriate care.

## Conclusion

In conclusion, this study highlights the significant advantages of fundus imaging with NIR trans-palpebral illumination for detecting and assessing choroidal nevi. This approach enhances nevus visibility and boundary definition compared to SLO. The close agreement between nevus basal diameter measurements in NIR fundus images and OCT B-scans confirms the accuracy and reliability of this technique for measurement of choroidal nevi. This method shows promise in enhancing clinical practice, improving the screening, diagnosis, and monitoring of choroidal nevi.

## Data Availability

All data produced in the present study are available upon reasonable request to the authors

## Reference

1. Shields JA, Shields CL. Intraocular tumors: an atlas and textbook: Lippincott Williams & Wilkins; 2008.

2. Shields CL, Lally SE, Dalvin LA, et al. White paper on ophthalmic imaging for choroidal nevus identification and transformation into melanoma. Translational Vision Science & Technology 2021;10:24–24.

3. Qiu M, Shields CL. Choroidal nevus in the United States adult population: racial disparities and associated factors in the National Health and Nutrition Examination Survey. Ophthalmology 2015;122:2071–2083.

4. Geiger F, Said S, Bajka A, et al. Assessing Choroidal Nevi, Melanomas and Indeterminate Melanocytic Lesions Using Multimodal Imaging—A Retrospective Chart Review. Current Oncology 2022;29:1018–1028.

5. Shields CL, Dalvin LA, Ancona-Lezama D, et al. Choroidal nevus imaging features in 3,806 cases and risk factors for transformation into melanoma in 2,355 cases: the 2020 Taylor R. Smith and Victor T. Curtin Lecture. Retina 2019;39:1840–1851.

6. Muftuoglu IK, Gaber R, Bartsch D-U, Meshi A, Goldbaum M, Freeman WR. Comparison of conventional color fundus photography and multicolor imaging in choroidal or retinal lesions. Graefe’s Archive for Clinical and Experimental Ophthalmology 2018;256:643–649.

7. Pe’er J, Sancho C, Cantu J, et al. Measurement of choroidal melanoma basal diameter by wide-angle digital fundus camera: a comparison with ultrasound measurement. Ophthalmologica 2006;220:194–197.

8. Vallabh N, Sahni J, Parkes C, Czanner G, Heimann H, Damato B. Near-infrared reflectance and autofluorescence imaging characteristics of choroidal nevi. Eye 2016;30:1593–1597.

9. Jonna G, Daniels AB. Enhanced depth imaging OCT of ultrasonographically flat choroidal nevi demonstrates 5 distinct patterns. Ophthalmology Retina 2019;3:270–277.

10. Torres VL, Brugnoni N, Kaiser PK, Singh AD. Optical coherence tomography enhanced depth imaging of choroidal tumors. American journal of ophthalmology 2011;151:586–593. e582.

11. Daniels AB, Veverka KK, Patel SN, Sculley L, Munn G, Pulido JS. Computing uveal melanoma basal diameters: a comparative analysis of several novel techniques with improved accuracy. International Journal of Retina and Vitreous 2019;5:1–10.

12. Shields CL, Kaliki S, Rojanaporn D, Ferenczy SR, Shields JA. Enhanced depth imaging optical coherence tomography of small choroidal melanoma: comparison with choroidal nevus. Archives of Ophthalmology 2012;130:850–856.

13. Yao X, Son T, Ma J. Developing portable widefield fundus camera for teleophthalmology: Technical challenges and potential solutions. Experimental Biology and Medicine 2022;247:289–299.

14. Rossi A, Rahimi M, Le D, et al. Portable widefield fundus camera with high dynamic range imaging capability. Biomedical Optics Express 2023;14:906–917.

15. Patel SN, Shi A, Wibbelsman TD, Klufas MA. Ultra-widefield retinal imaging: an update on recent advances. Therapeutic advances in ophthalmology 2020;12:2515841419899495.

16. Toslak D, Thapa D, Chen Y, Erol MK, Chan RP, Yao X. Trans-palpebral illumination: an approach for wide-angle fundus photography without the need for pupil dilation. Optics letters 2016;41:2688.

17. Son T, Ma J, Toslak D, et al. Light color efficiency-balanced trans-palpebral illumination for widefield fundus photography of the retina and choroid. Scientific Reports 2022;12:13850.

18. Rahimi M, Rossi A, Son T, Yao X. High dynamic range fundus camera with trans-palpebral illumination. Investigative Ophthalmology & Visual Science 2023;64:5016–5016.

19. Panwar N, Huang P, Lee J, et al. Fundus photography in the 21st century—a review of recent technological advances and their implications for worldwide healthcare. Telemedicine and e-Health 2016;22:198–208.

20. Rossi A, Rahimi M, Son T, Chan RP, Heiferman MJ, Yao X. Preserving polarization maintaining photons for enhanced contrast imaging of the retina. Biomedical Optics Express 2023;14:5932–5945.

21. Feng X, Yu Y, Zou D, et al. Functional imaging of human retina using integrated multispectral and laser speckle contrast imaging. Journal of biophotonics 2022;15:e202100285.

22. Huang Z, Jiang Z, Hu Y, et al. Retinal choroidal vessel imaging based on multi-wavelength fundus imaging with the guidance of optical coherence tomography. Biomedical Optics Express 2020;11:5212.

23. Francis JH, Pang CE, Abramson DH, et al. Swept-source optical coherence tomography features of choroidal nevi. American Journal of Ophthalmology 2015;159:169–176. e161.

24. Alterini T, Díaz-Doutón F, Burgos-Fernández FJ, González L, Mateo C, Vilaseca M. Fast visible and extended near-infrared multispectral fundus camera. Journal of biomedical optics 2019;24:096007–096007.

25. de Carvalho ER, Hoveling RJ, van Noorden CJ, Schlingemann RO, Aalders MC. Functional imaging of the ocular fundus using an 8-band retinal multispectral imaging system. Instruments 2020;4:12.

26. Burgos-Fernández FJ, Alterini T, Díaz-Doutón F, et al. Reflectance evaluation of eye fundus structures with a visible and near-infrared multispectral camera. Biomedical optics express 2022;13:3504–3519.

27. Venkatesh R, Pereira A, Sangai S, et al. Variability in imaging findings in choroidal nevus using multicolor imaging vis-à-vis color fundus photography. Journal of Current Ophthalmology 2020;32:285.

28. Azzolini C, Di Nicola M, Pozzo Giuffrida F, et al. Retromode Scanning Laser Ophthalmoscopy for Choroidal Nevi: A Preliminary Study. Life 2023;13:1253.

29. Xu Y, Lou L, Wang Y, et al. Epidemiological study of uveal melanoma from US surveillance, epidemiology, and end results program (2010–2015). Journal of Ophthalmology 2020;2020.

30. Standardization IOf. Ophthalmic Instruments-Fundus Cameras-International Standard, ISO 10940. 2007.

31. Rahimi M, Rossi A, Son T, et al. Evaluating spatial dependency of the spectral efficiency in trans-palpebral illumination for widefield fundus photography. Biomedical Optics Express 2023;14:5629–5641.

32. Toslak D, Son T, Erol MK, et al. Portable ultra-widefield fundus camera for multispectral imaging of the retina and choroid. Biomedical optics express 2020;11:6281.

33. Sanchez-Brea LM, Quiroga JA, Garcia-Botella A, Bernabeu E. Histogram-based method for contrast measurement. Applied optics 2000;39:4098–4106.

34. Ma F, Yuan M, Kozak I. Multispectral Imaging (MSI): Review of Current Applications. Survey of Ophthalmology 2023.

35. Bird AC. Therapeutic targets in age-related macular disease. The Journal of clinical investigation 2010;120:3033–3041.

